# State and County-Level Factors Associated with the Effectiveness of Stay-At-Home Orders Issued in the United States in Response to COVID-19

**DOI:** 10.1101/2023.04.27.23289229

**Authors:** Forrest DL Barker, Ali Mirzadadeh, Neelam Sekhri Feachem

## Abstract

**Title:** State and County-Level Factors Associated with the Effectiveness of Stay-At-Home Orders Issued in the United States in Response to COVID-19

**Background:** To slow the spread of COVID-19 and protect medical facilities from overflowing, Stay-At-Home Orders (SAHOs) were issued in the United States during the spring of 2020. These orders had variable levels of effectiveness and profound consequences that continue to manifest long after their termination. This study aimed to assess if state and county-level population characteristics could explain variability in SAHO effectiveness as measured by the effective reproductive number (R_t_).

**Methods:** We calculated the R_t_ for the 40 states which enacted SAHOs, and also for a sample of 289 counties that issued SAHOs in 2020, using EpiEstim R Package based on the states’ and counties’ daily case data. We determined SAHOs to be effective if, three weeks after their implementation, R_t_ was equal to or less than one. Wilcoxon rank sum tests and logistic regression were used to determine if population characteristics (age, income, level of education, political orientation, percent of non-English speaking people, racial and ethnic compositions), and percentage of frontline workers, percent of eligible people vaccinated by July 2021, level of viral transmission, and other Non-Pharmaceutical-Interventions (NPI) enacted before the SAHO, were associated with effectiveness of SAHOs.

**Results:** SAHOs were effective in 20 (50%) states. No significant differences were found in the characteristics studied between states with effective and ineffective SAHOs. SAHOs were effective in 54% of counties. Counties with effective SAHOs had fewer days of NPIs before the SAHOs in comparison to counties with ineffective SAHOs (Median 24 vs. 34, p-value 0.005). All other characteristics considered showed non-significant differences. In multivariate analysis, days of NPIs before the SAHOs remained the only significant factor for effective SAHOs in studied counties.

**Conclusion:** Our analysis suggests that SAHO effectiveness may be influenced by the implementation of prior public health interventions but is not likely to be related to the other characteristics studied. These findings should be considered when assessing when and how to implement SAHOs in future epidemics to limit the spread of an infectious respiratory disease.

## INTRODUCTION

In early December 2019, a cluster of idiopathic pneumonia cases was reported in Wuhan, China.^1^ These are thought to be the first identified cases of a disease called COVID-19 caused by a novel coronavirus named Severe Acute Respiratory Coronavirus Syndrome-2 (SARS-CoV-2) or colloquially the COVID-19 virus.^2^ By the end of January 2020, COVID-19 was classified as a public health emergency of international concern and cases had been detected in 21 countries spread across Asia, Europe, North America, and Oceania. A few weeks later, the World Health Organization (WHO) declared COVID-19 a global pandemic.^3^ As of April 1, 2023, the WHO had globally recorded 761,402,282 confirmed cases and 6,887,000 confirmed deaths, making COVID-19 one of the most severe pandemics in history.^4^ In the United States (U.S.) alone the COVID-19 virus has infected over 100 million individuals, more than one million of whom have died.^5^

The number of active COVID-19 cases has periodically spiked, causing hospitals to reach and surpass their capacity. This in turn has forced care and resources to be rationed.^6–10^ For example, medical facilities in the U.S. have, at times, needed to suspend non-emergent services to prioritize resources for overflowing emergency rooms and intensive care units (ICUs). At the same time medical supply chains have been disrupted by excessive demand and supply shortages.^11–13^ This was particularly true at the start of the pandemic when treatments for COVID-19 were unrefined and vaccines were unavailable, leaving populations particularly vulnerable. The lack of pharmaceutical protection forced nations to implement Non-Pharmaceutical Interventions (NPI’s) to reduce transmission of COVID-19 and avert the collapse of their medical systems. In the U.S. this effort was termed “flattening the curve”.^14^ Without such measures, hospitals would overflow, and patients would be left untreated, drastically increasing all-cause mortality.

Throughout the pandemic many NPI’s have been utilized to reduce transmission with varying levels of success.^15,16^ Common NPIs have include mask wearing, quarantine (confinement of individuals who may be infected), isolation (confinement of individuals who are infected), restricting travel, prohibiting gatherings, requiring individuals to maintain distance between each other, closing schools, closing non-essential businesses, and full lockdowns or Stay-at-Home Orders (SAHOs). Implementation of these NPIs has come at the cost of reducing personal liberties and economic activity, increasing inequality, and a myriad of unintended public health side effects such as increased consumption of drugs and alcohol, higher rates of mental health disorders, and rising rates of child abuse and intimate partner violence.^17–22^

Prior to COVID-19, the efficacy and consequences of NPIs were poorly researched and largely based on theoretical knowledge.^23^ However, throughout the pandemic, countries have implemented and relaxed various NPI’s in response to rising and falling infection rates. This has created a natural experiment which allows for real world assessment of the efficacy of interventions. As a result, an enormous amount of research on the subject has been conducted in the last few years. Assessments have utilized numerous methodologies including mathematical models, surveys, opinions, direct observations and measurements, and indirect observations via proxy data. In addition to the multitude of methodologies for assessing NPI effectiveness, there is substantial variability in the metrics used for quantification, making it challenging to compare analyses. In general, it has been found that effectiveness of NPIs varies between locations and that multiple interventions have stronger effects than any single intervention.^23,24^

SAHOs, which inherently include other NPI’s, have been found to be very effective at reducing transmission of COVID-19. However, it is also estimated that SAHOs in the US resulted in up to $64 billion a week in lost revenue and brought a grim set of unintended consequences.^21,22,25–28^ Often described as a blunt instrument or last resort, the far-reaching impacts of SAHOs warrant an in-depth investigation into the state and county-level factors that influence their efficacy. A number of investigations have shown that SAHO’s are effective at reducing population mobility and that SAHO’s are associated with a decrease in transmission and mortality at a state and county level.^29^ However, it has not been determined if certain population characteristics are associated with more or less effective SAHO’s. This paper investigates whether various state or county level factors could have influenced the efficacy of SAHOs issued in the U.S. during the spring of 2020. The threat of a future pandemic requiring lockdowns has drawn the attention of the world. Information presented here may help inform how future SAHOs can be coordinated in a manner that maximizes their impact while minimizing their duration and harm.

## MATERIALS AND METHODS

We performed an independent cross-sectional analysis looking for differences in communities with effective and ineffective SAHOs during the spring of 2020. Effective SAHOs were defined by communities achieving a local R_t_ (i.e., basic reproductive number at a point in time to measure transmissibility of the virus), of ≤ 1 after three weeks of a SAHO. Ineffective SAHOs were defined by having an R_t_ > 1 three weeks after order implementation. We initially proposed quantifying SAHO effectiveness as a continuous variable defined by the change in R_t_ from the first week of SAHO implementation to the third week of the SAHO, however, we found this definition favored communities with higher initial values of R_t_ and did not accurately describe whether a SAHO accomplished the goal of reducing the number of cases in a community.

Factors we assessed for possibly influencing SAHO effectiveness included:

● Percentage of essential workers
● R_t_ when order was enacted.
● Time between first case and SAHO enactment
● Average population age
● Average population income
● Political affiliation
● Average educational attainment
● Other NPI orders in place prior to and during SAHO
● Cumulative days of other NPI orders prior to SAHO
● Percentage of non-English speaking individuals
● Percentage of individuals identifying as African American
● Percentage of individuals identifying as Hispanic

We originally proposed to also study whether community mobility influenced SAHO effectiveness. However, this analysis was conducted and published by Wellenius et al. while we were conducting our investigation.^30^ As the influence of mobility on SAHOs was no longer a novel investigation we removed it from our list of variables, to prioritize others.

All 40 states that issued SAHOs were included in this sample. For county level analysis a sample of 429 counties was obtained from the American Community Survey (ACS). This sample is designed to be representative of the population of the US. The sample was filtered to remove counties without SAHOs during the study period, where there were too few cases of COVID-19 for accurate calculations of R_t_, and where the first case was not reported until after the start of the SAHO. This filtering yielded a final sample of 289 counties spread across the U.S.

The reproductive number R_t_ was calculated based on confirmed cases of COVID-19 reported in the Johns Hopkins University Coronavirus Resource Center Database.^31^ This dataset included cumulative daily anonymized COVID-19 cases aggregated by county. New daily cases were defined as the change in cumulative cases from one day to the next. Dates with negative total daily cases were adjusted to zero cases for that day. For state level analysis, the daily case data for all counties in each state were summed. An attempt was made to remove cases known to have occurred in congregate living settings such as prisons and nursing homes. However, available data on these cases were insufficient to make a robust adjustment. R version 4.0.2. and R Studio were used for organization and manipulation of case data. EpiEstim R Package was then used to calculate R_t_ for states and counties based on daily case data. A serial interval (SI) of 5.2 days with a standard deviation of 0.7 days, as reported in the meta-analysis by Alene et al, was used in calculating R_t_.^32^ For each state and county, R_t_ was measured at two time points, the week following the start of the SAHO and the week 21 to 27 days following the start of the SAHO. These time points are estimated to represent viral reproductive number during the week preceding the SAHO implementation and two weeks post implementation, accounting for the 6.5 day mean incubation period also reported by Alene et al.^32^ R_t_ was calculated as an average over a seven-day time period to account for lower case reporting on weekends compared to weekdays. Locations where fewer than ten cases were reported during either of the seven-day periods used for calculating R_t_ were excluded as so few cases did not allow for accurate calculation of R_t_. Additionally, calculated R_t_ values that were greater than 2 standard deviations above the mean were excluded as outliers. These outliers were thought to represent either an outbreak in a congregate setting, or a batch of previously unreported cases and thus not representative of community transmission.

Dates of public health orders including the start of SAHOs were retrieved from the CUSP database maintained by Boston University.^33^ For each location, we identified NPI’s that preceded SAHO implementation. The number of days each NPI was in place prior to the SAHO were then summed to give a cumulative number of days with prior NPI orders. For example, if a location had one order that began ten days before the SAHO and another began five days before the SAHO, that location would be classified as having 15 days of prior NPI orders. The orders included in this measurement were: declaring a state of emergency, school closures, business closures, and prevention of large gatherings.

The IPUMS database was used to access and organize a five-year sample (2015-2019) of the census bureau’s ACS community survey, which is designed to be 5% of the U.S. population. This sample contained over 15.9 million unique responses including data on individuals’ sex, income, age, race, ethnicity, occupational industry, ability to speak English, educational attainment, county and state of residence, and the weight each response should carry to represent the state population. Responses were filtered to exclude entries where county of residence was not reported. Data were then aggregated at both the state and county level and further filtered to only include locations with SAHOs and a sufficient number of reported cases. The final sample contained 5,787,898 responses across 289 counties, with responses from a mean of 2.8% of the population in each county (1.5% minimum and 5% maximum).

For each county and state/territory, the mean value was calculated for age and years of education. Years of education were coded from 0-11 where 0 represents no schooling, 1 represents schooling up to fourth grade, 2 represents schooling up to eighth grade, 3-10 each represent one additional grade or year of schooling above eighth grade, 11 represents five or more years of college. The median value was calculated for income as these data had outliers with high earnings. Income data were also altered by the ACS survey which reports incomes over $999,999 as $999,999. This is done to protect anonymity of respondents. The percentage of respondents who reported speaking little to no English was calculated and used to estimate percent of non-English speaking individuals. Similarly, the percentage of respondents who identified as Hispanic, and who identified as African American were calculated.

In the ACS survey, industry is logged as a 5-digit code based on the North American Industrial Classification System (NAICS). We identified 22 NAICS codes that contained most front-line workers including: essential consumer business such as grocery stores, transportation workers, first responders, and medical practitioners. The codes included were 4470, 4971, 4972, 5070, 5391, 6080, 6180, 6170, 6390, 6370, 7690, 7970, 8090, 8170, 8180, 8191, 8192, 8270, 8290, 8370, 8380, and 8470. The percentage of respondents who reported working in one of these industries was then calculated as an estimate for the community’s percent of front-line workers. We identified front line workers based on industry rather than occupation because industry has a larger influence on whether someone is a front-line worker. For example, a manager at a grocery store is a front-line worker while a manager of a software company is not. In this example the occupation of manager is not indicative of whether a job is front-line while industry, in this case, grocery store or software development, is.

The ACS survey lists response weights which indicate how many people in the population each response represents. These weights are based on state population data. Weighting was utilized for state level calculations but not for county level calculations. The unweighted calculations at the county level allowed us to capture intra-state variability better than the weighted sample.

Data on vaccination rates in each location was collected from the U.S. Centers for Disease Control and Prevention’s COVID-19 Data Tracker.^5^ Data on the percentage of people who voted for Trump in 2020 was obtained from MIT’s Election Lab Database.^34^

Utilizing R Version 4.0.2 and R Studio, we ran Wilcoxon rank sum tests with all the above-mentioned variables to compare their distribution in populations with successful or unsuccessful SAHOs. We utilized an alpha value of 0.05 as our rejection criteria. The Wilcoxon test was utilized for hypothesis testing because data on the variables did not have a normal distribution or contained outliers which could not be excluded as they represented valuable data points on unique communities. We then created logistic regression models controlling for each of the 12 factors assessed to determine if any factors were functioning as confounders or effect modifiers of other factors. We chose to classify variables as having a significant effect if their presence in the logistic regression model reduced the p-value of other variables by more than 10%.

## RESULTS

A summary of the sample of ACS data used for our analysis is shown in Table 1. Variables that were not from the ACS survey, such as percent of eligible population vaccinated, were derived from the whole population rather than a sample. Thus, the demographics of these population-based variables are not detailed.

**Table 1:**
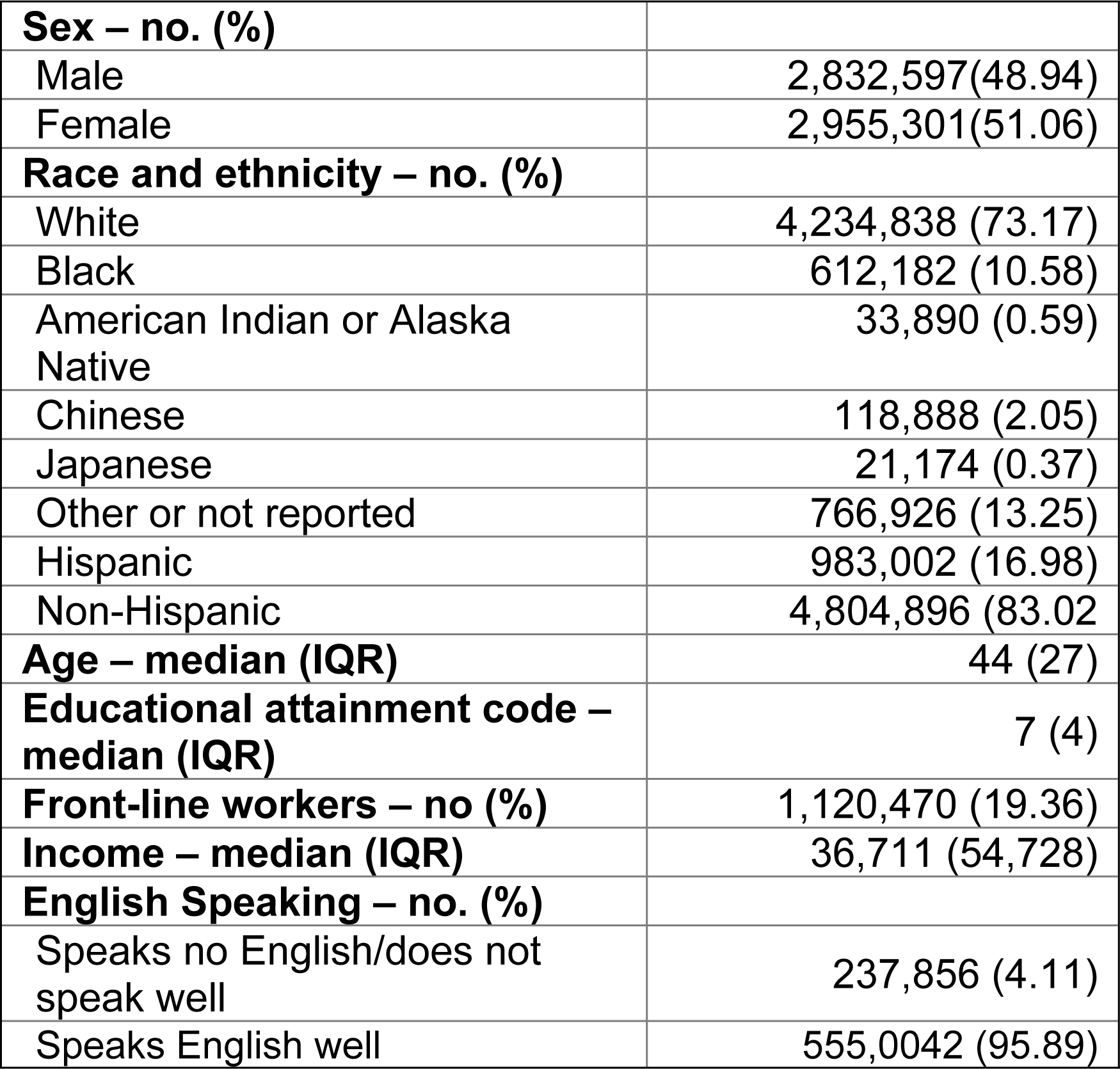
ACS survey sample responses (n=5,787,898)

Figure 1 shows the locations of the counties which made up our sample for analysis at the county level.

**Figure 1:**
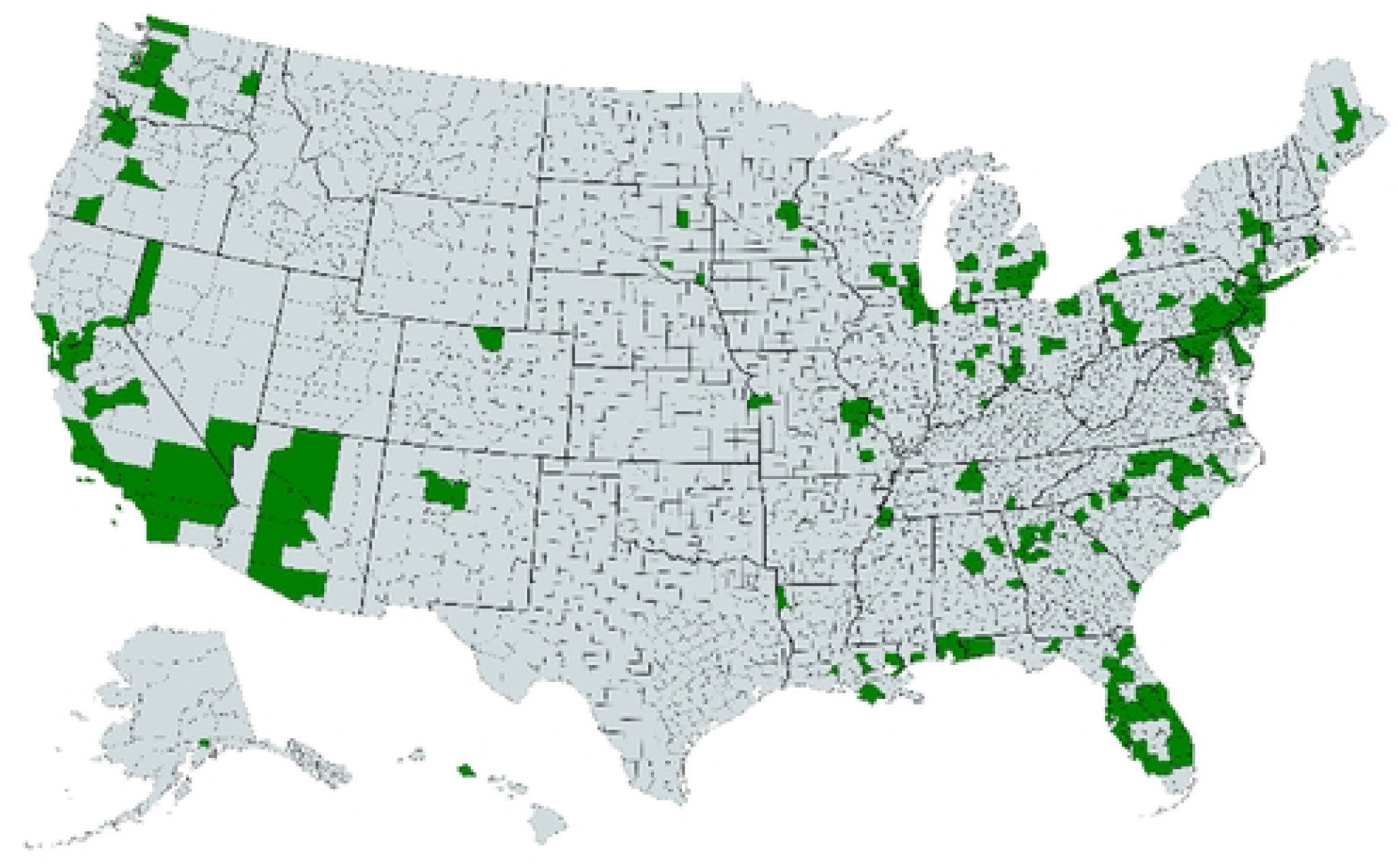
Counties included in analysis n=289.

**(See Separate File for All Figures)**

For state level analysis, we first grouped states into effective or ineffective SAHOs. States that achieved an R_t_ value of one or lower after three weeks of SAHO were said to have effective orders while states that did not achieve an R_t_ value of less than one during the same time period were classified as having ineffective SAHOs. Based on these criteria it was found that, of the 40 states with SAHOs, 50% were successful. Table 2 shows the variation of characteristics we assessed for states with effective and ineffective SAHOs.

**Table 2:** Difference in state level characteristics by SAHO effectiveness.

|  | Effective (n=20) |  | Non-effective (n=20) |  | Wilcoxon rank sum test |  |
| --- | --- | --- | --- | --- | --- | --- |
|  | Median | IQR | Median | IQR | W | p-value |
| Time between first case and SAHO start (days) | 18.5 | 9.5 | 23 | 7.5 | 254.5 | 0.144 |
| Rt at Start of SAHO | 1.96 | 0.99 | 1.96 | 1.26 | 179 | 0.583 |
| Cumulative days of NPI orders (days) | 26 | 14.8 | 30 | 14.5 | 226.5 | 0.311 |
| Front-line workers (%) | 20.0 | 2.3 | 20.8 | 2.1 | 209 | 0.138 |
| Educational attainment | 7.70 | 0.31 | 7.74 | 0.43 | 164 | 0.950 |
| Non-English speaking (%) | 1.62 | 2.24 | 1.94 | 1.88 | 163 | 0.975 |
| Income (2019 USD) | 37,859 | 6,366 | 38,300 | 5,088 | 149 | 0.704 |
| Age (years) | 44 | 1 | 44 | 1.5 | 184.5 | 0.461 |
| Eligible population vaccinated (%) | 68.6 | 15.7 | 62.9 | 18.5 | 192.5 | 0.850 |
| Voted for Trump in 2020 presidential election (%) | 45.4 | 14.7 | 48.8 | 12.8 | 216 | 0.478 |
| Identify as Hispanic (%) | 8.35 | 2.70 | 7.24 | 7.34 | 143 | 0.573 |
| Identify as African American (%) | 11.9 | 12.7 | 10.4 | 10.9 | 164 | 0.950 |

As with states, our sample of counties was split into two groups, those with effective SAHOs and those with ineffective SAHOs. Effectiveness was determined based on the same criteria as states. Of the 289 counties in our sample, 158 (54%) were determined to be successful. Table 3 shows the variation in characteristics of counties with successful SAHOs and those with unsuccessful SAHOs.

**Table 3:** Difference in county level characteristics by SAHO effectiveness.

|  | Effective (n=185) |  | Non-effective (n=127) |  | Wilcoxon rank sum test |  |
| --- | --- | --- | --- | --- | --- | --- |
|  | Median | IQR | Median | IQR | W | p-value |
| Time between first case and SAHO start (days) | 14 | 8.8 | 15 | 9.5 | 10678 | 0.642 |
| Rt at Start of SAHO | 2.07 | 1.57 | 1.91 | 1.36 | 9494 | 0.227 |
| Cumulative days of NPI orders (days) | 24 | 22 | 34 | 20 | 12044 | **0.005** |
| Front-line workers (%) | 19.9 | 3.6 | 20.0 | 3.8 | 10264 | 0.905 |
| Educational attainment | 7.78 | 0.64 | 7.72 | 0.68 | 9539 | 0.252 |
| Non-English speaking (%) | 1.27 | 1.72 | 1.47 | 1.92 | 10810 | 0.515 |
| Income (2019 USD) | 37,769 | 10,384 | 36,652 | 9,766 | 9294 | 0.136 |
| Age (years) | 45 | 4 | 45 | 3 | 9207 | 0.104 |
| Eligible population vaccinated (%) | 54.6 | 13.8 | 51.5 | 17.0 | 9111.5 | 0.080 |
| Voted for Trump in 2020 presidential election (%) | 44.8 | 23.5 | 45.9 | 20.5 | 10300 | 0.895 |
| Identify as Hispanic (%) | 6.40 | 7.26 | 5.94 | 6.99 | 9864 | 0.493 |
| Identify as African American (%) | 7.43 | 13.1 | 7.77 | 12.1 | 11143 | 0.262 |

The characteristics assessed were factors or proxies for factors we hypothesized could have influenced how well populations complied with SAHOs, and how effective SAHOs were at reducing transmission of COVID-19. Wilcoxon rank sum tests were utilized to test the null hypothesis that there was not a difference in the distribution of characteristics between communities with successful versus unsuccessful SAHOs. We utilized an alpha value of 0.05 as the rejection criteria.

Figures 2 through 13 show the distribution of data points at the state and county levels. Each graph shows how a characteristic varied among locations with successful SAHOs (green) and unsuccessful SAHOs (red). For clarity, data points on these graphs have been binned into 30 distinct groups evenly distributed along the Y axis.

### • Days between first case and SAHO start

The first factor we considered was the number of days between the first case and the start of a SAHO. We suspected that as this time period grew, so would the number of undetected cases of COVID-19. Greater numbers of individuals with undetected cases could lead to increased household transmission once SAHOs were put in place, causing them to be less effective at reducing R_t_. However, we did not find a difference in the distribution of this time period between communities (states or counties) with effective versus ineffective SAHOS (p=0.144 states; p= 0.642counties).

**Figure 2:**
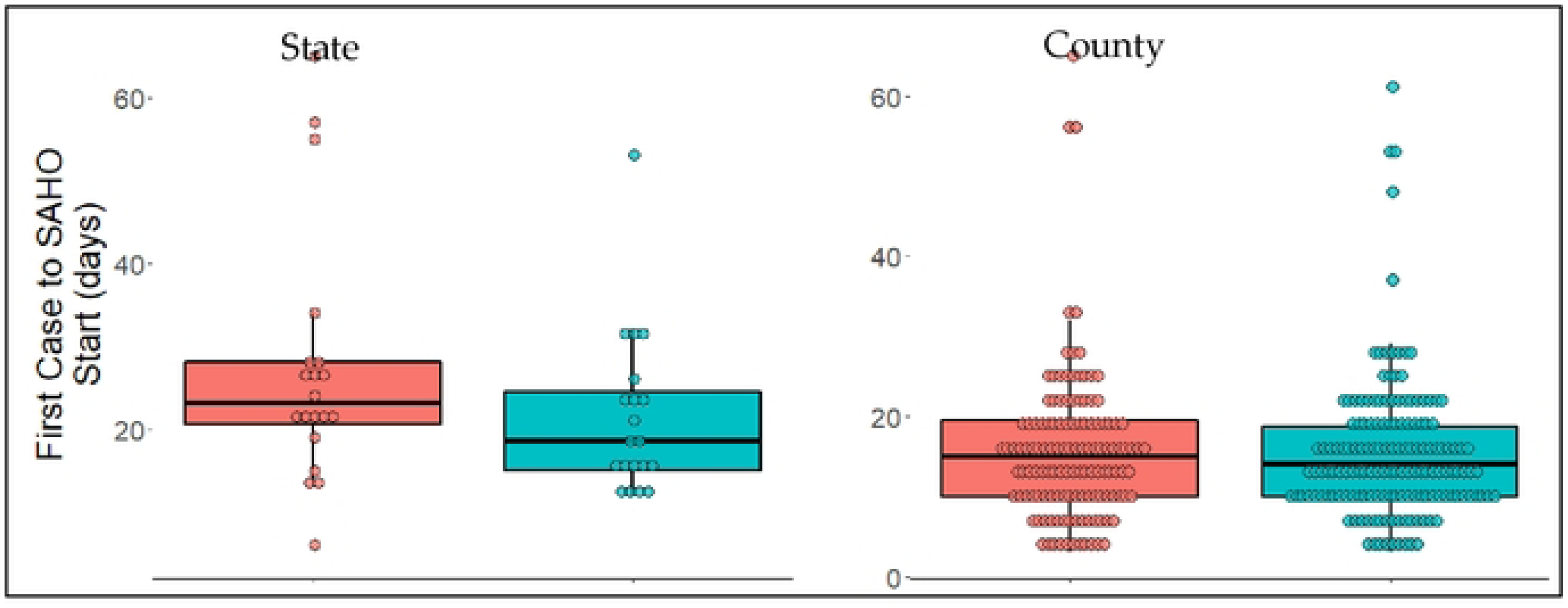
Days between first case and SAHO Start. Non-successful SAHOs are shown on the left in red, successful SAHOs are shown on the right in green.

### • R_t_ value at start of SAHO

We then considered the viral reproductive value at the start of SAHOs. Other studies have reported that NPIs, including SAHOs, can reduce R_t_ by finite amounts which would make it easier for SAHOs to be effective in communities with lower initial R_t_ values. Comparison of R_t_ distribution between groups yielded non-significant results (p=0.583 states; p=0.227 counties). Thus, we could not reject the null hypothesis that the distribution of initial viral reproduction values was the same for communities with effective versus ineffective SAHOs.

**Figure 3:**
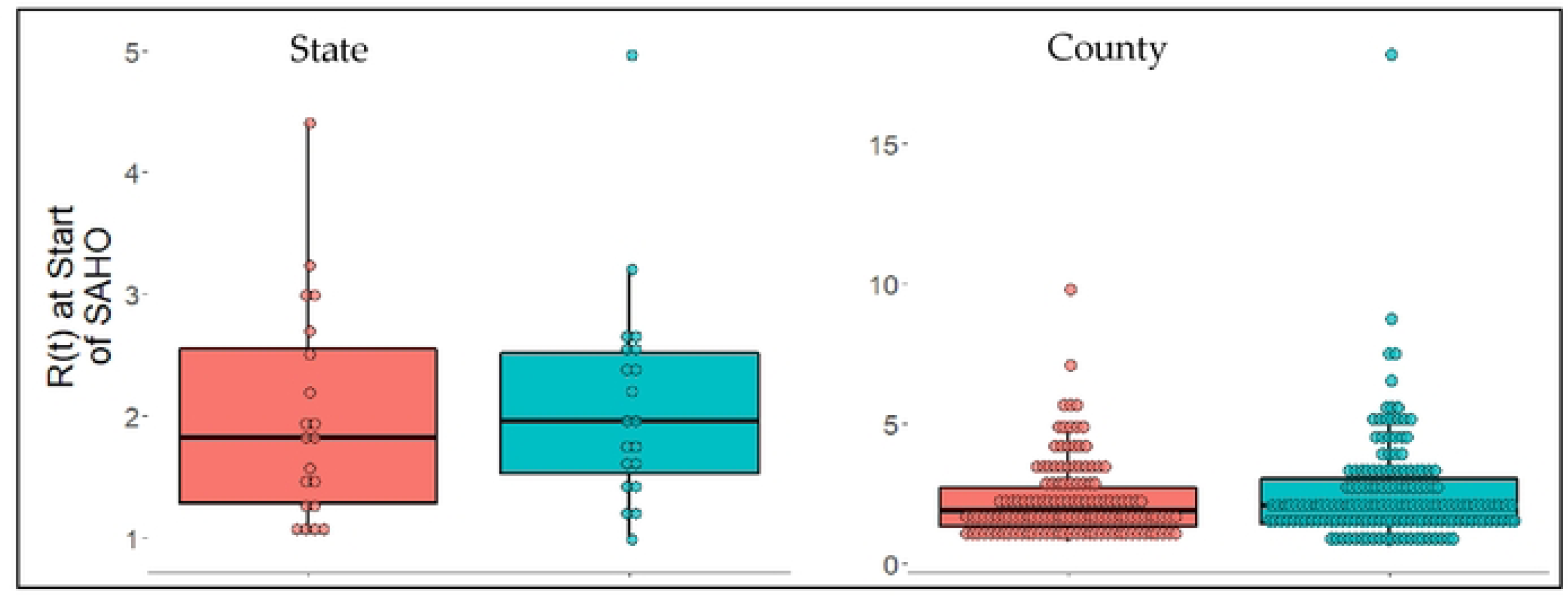
R_t_ calculated for the first week of SAHO. Non-successful SAHOs are shown on the left in red, successful SAHOs are shown on the right in green.

### • Cumulative days of NPI orders prior to SAHO start

After looking at initial R_t_ values, we assessed if the cumulative number of days of alternate NPI orders, before SAHOs were implemented could have influenced effectiveness. Other studies have reported that multiple NPI orders were more effective than a single NPI order. Based on this we expected that a greater number of days of other NPI orders before a SAHO began would result in a SAHO being more likely to drop the R_t_ value to below 1. When we tested this hypothesis at the state level, we did not find a difference in the distribution of days of prior NPI orders between effective and ineffective SAHOs (p=0.311). However, we found a significant difference between these groups of SAHOs at the county level (p=0.005). Surprisingly this difference was not in our hypothesized direction. Rather we found that the median number of days of prior NPI orders in counties with effective SAHOs was lower than the median in counties with ineffective SAHOs.

**Figure 4:**
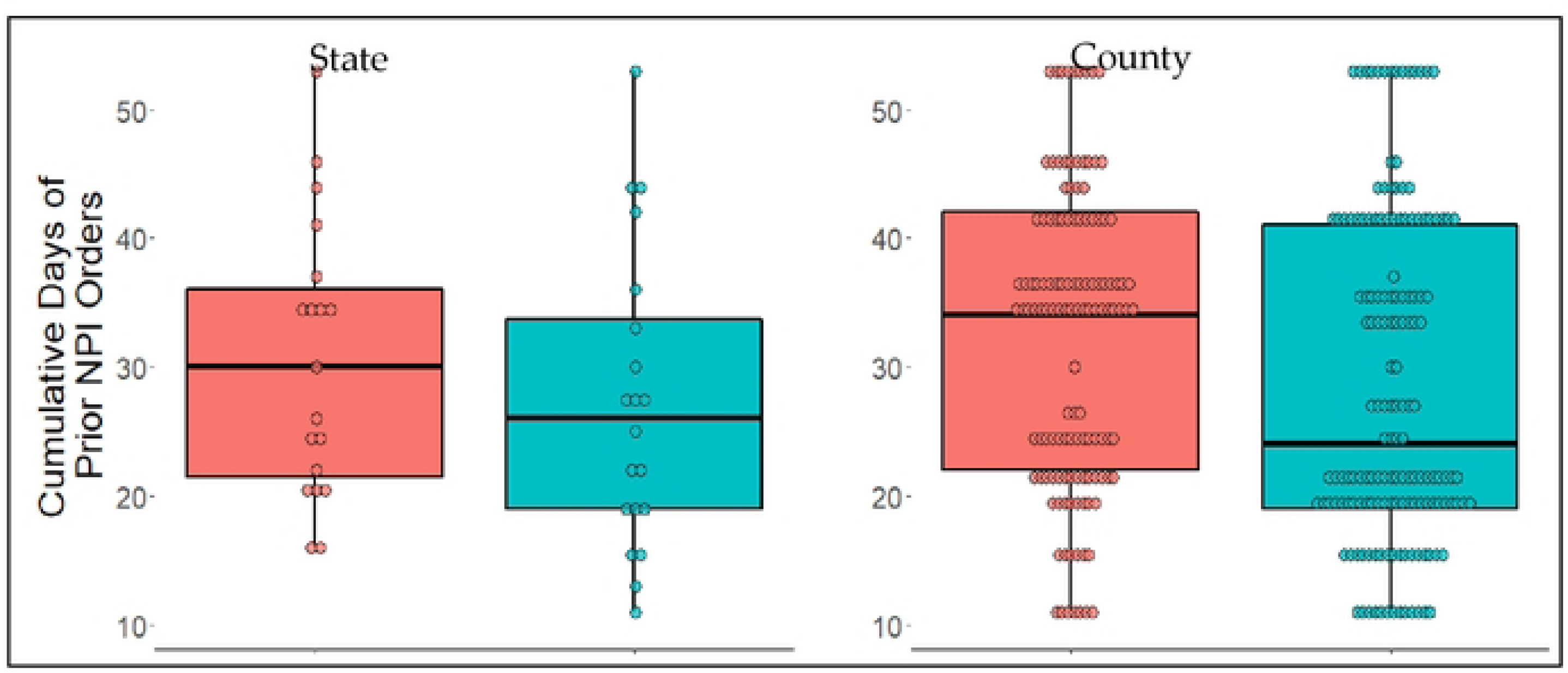
The cumulative number of days other NPI orders were in place before the start of a SAHO. Non-successful SAHOs are shown on the left in red, successful SAHOs are shown on the right in green.

### • Percent of front-line workers

The next characteristic we considered was the percent of front-line workers. The jobs of these workers were not affected by SAHOs, and they continued to have high levels of interaction and contact with other community members. Thus, we expected that the effectiveness of SAHOs would be reduced in communities with a greater proportion of front-line workers. Our analysis, however, did not find a difference in SAHO effectiveness for either states or counties based on the percent of front-line workers (p=0.138 states; p=0.905 counties).

**Figure 5:**
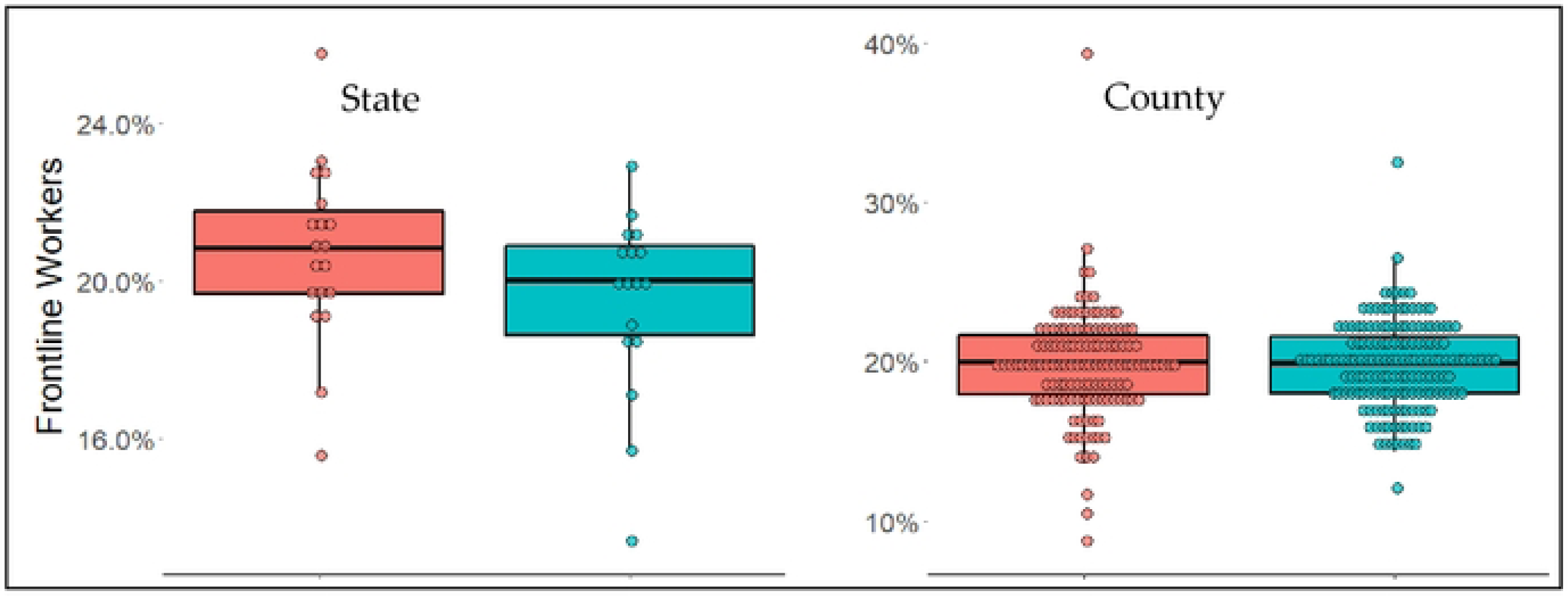
The percent of ACS survey respondents who reported working in industries identified as front-line. Non-successful SAHOs are shown on the left in red, successful SAHOs are shown on the right in green.

### • Educational attainment

Educational attainment has commonly been associated with behavioral differences. For example, studies have found correlations between college education and political affiliations. We hypothesized that the level of education in a community could influence how that community responds to public health orders. We tested the null hypothesis that there was no difference in distribution of educational attainment between communities with successful and unsuccessful SAHOs and found that there was not a significant difference (p=0.950 states; p=0.252 counties).

**Figure 6:**
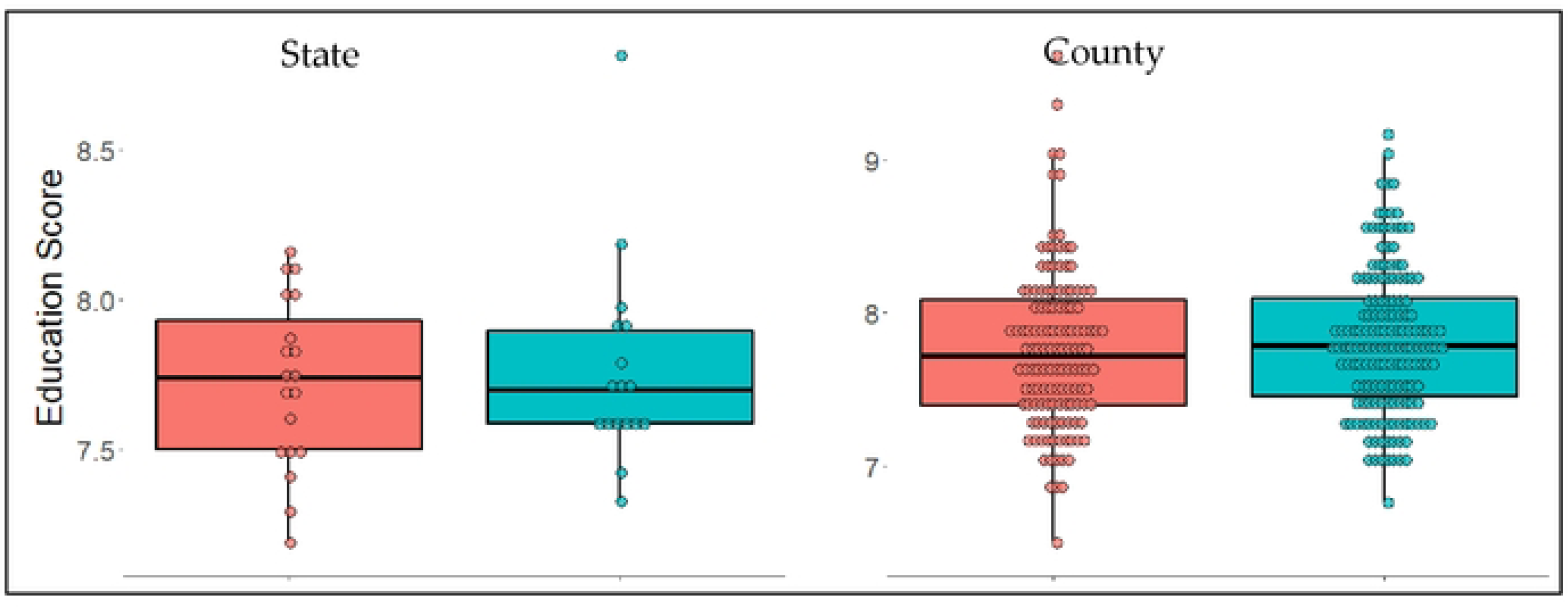
Educational scores relating to the highest level of education attained by respondents of ACS survey. On this scale 0 represents no schooling and 11 represents 5+ years of college. Non-successful SAHOs are shown on the left in red, successful SAHOs are shown on the right in green.

### • Non-English-speaking population

Public health messaging in the US was largely carried out in English. This messaging spread awareness of SAHOs and gave explanations of their purpose, goals, and guidelines. It is possible that non-English speaking individuals were not exposed to this messaging, which could have altered their behavior while SAHOs were in effect. We assessed if the proportion of individuals in states or counties who spoke little to no English differed between effective and ineffective SAHOs. Our tests did not find a significant difference in English speaking abilities based on the successfulness of SAHOs (p=0.975 states; p=0.515 counties).

**Figure 7:**
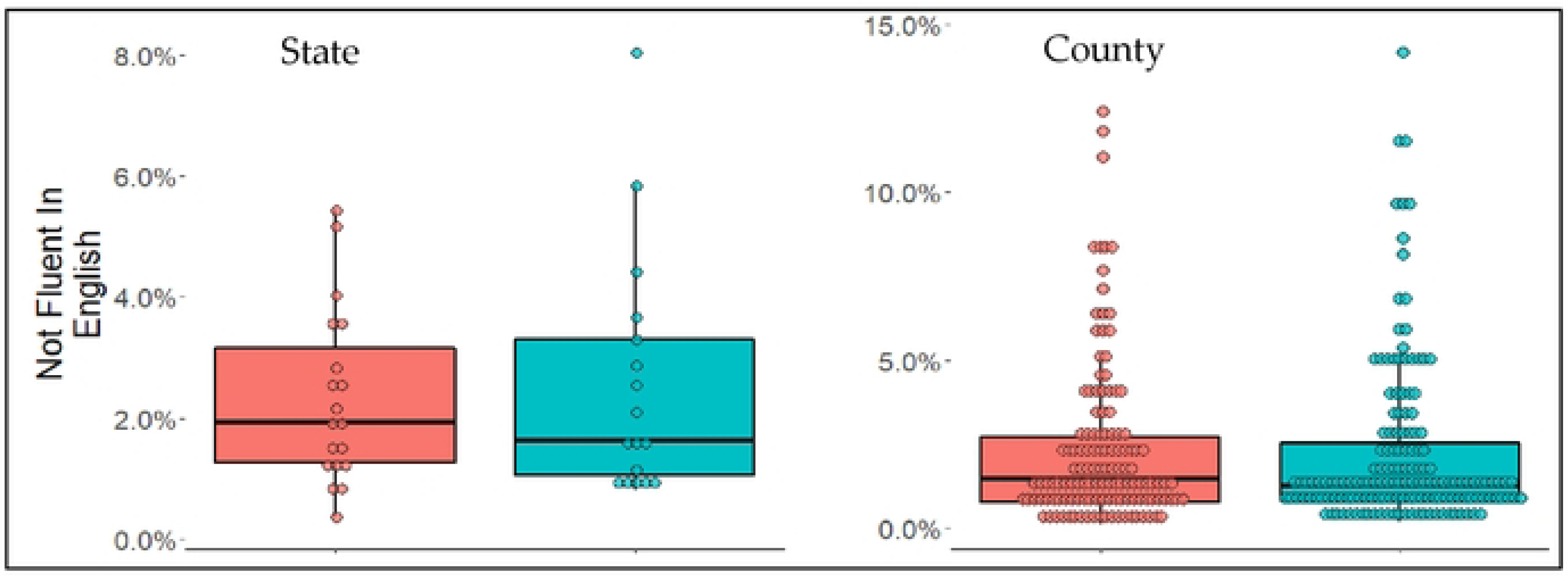
The percent of ACS survey participants who reported they did not speak English or did not speak English well. Non-successful SAHOs are shown on the left in red, successful SAHOs are shown on the right in green.

### • Income

The financial consequences of SAHOs issued in spring of 2020 were enormous. Compliance with SAHOs for many individuals meant loss of income and financial security while the businesses where they worked were closed or operating at reduced capacity. Additionally, many individuals and families faced large costs associated with working and schooling from home. To assess whether financial security could have influenced compliance with SAHOs thereby altering their effectiveness, we looked for a difference in the distribution of median incomes in states and counties with successful versus unsuccessful SAHOs. These tests yielded insignificant results, suggesting that the distribution of income is similar between the two groups at both the state and county level (p=0.704 states; p=0.136 counties).

**Figure 8:**
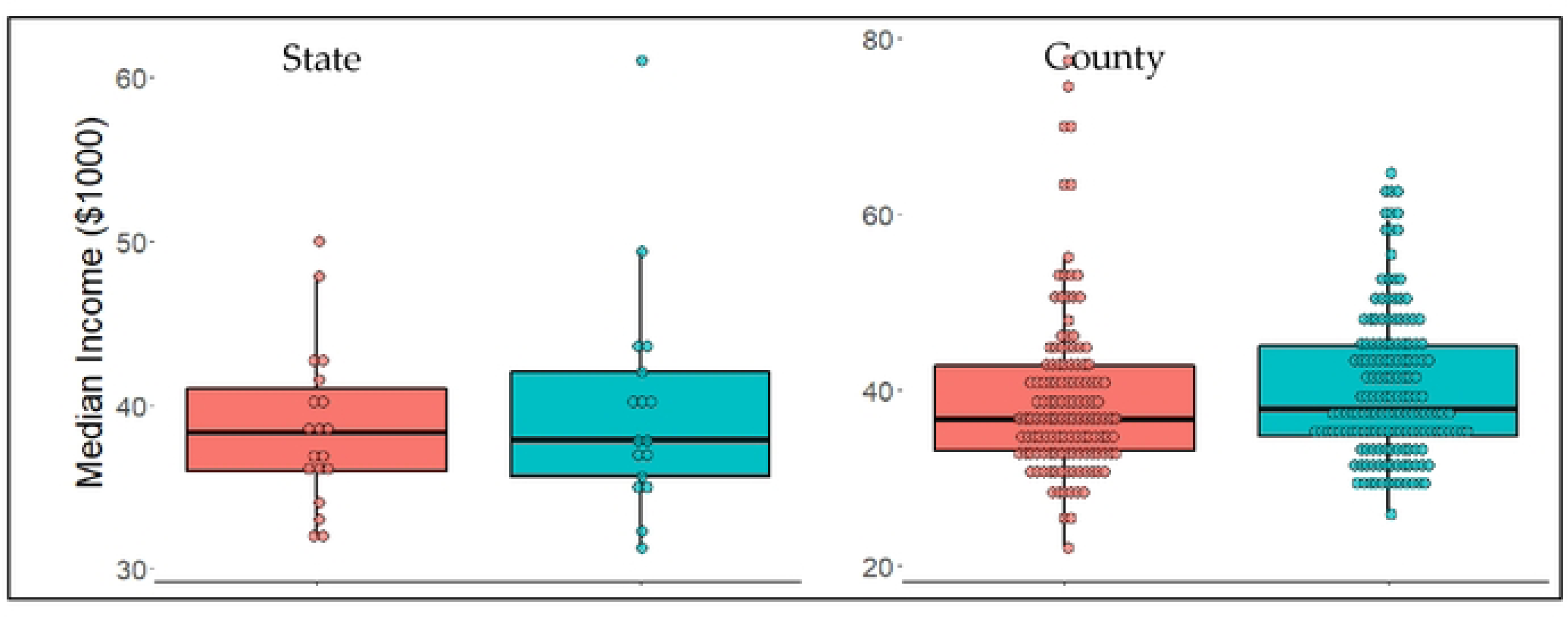
Median income in thousands of US Dollars reported in ACS survey responses (adjusted for inflation to match 2019 USD value). Unsuccessful SAHOs are shown on the left in red, successful SAHOs are shown on the right in green.

### • Age

Age has a profound impact on individuals’ behavior including mobility and social practices. Studies have found that SAHO effectiveness is negatively correlated with mobility and number of social interactions. Given this finding we expected that the median age of a population could influence whether a SAHO could reduce the viral reproductive value to less than one after three weeks. Contrary to our expectation, analysis did not show any difference in the median age of populations based on SAHO successfulness (p=0.461 states; p=0.104 counties).

**Figure 9:**
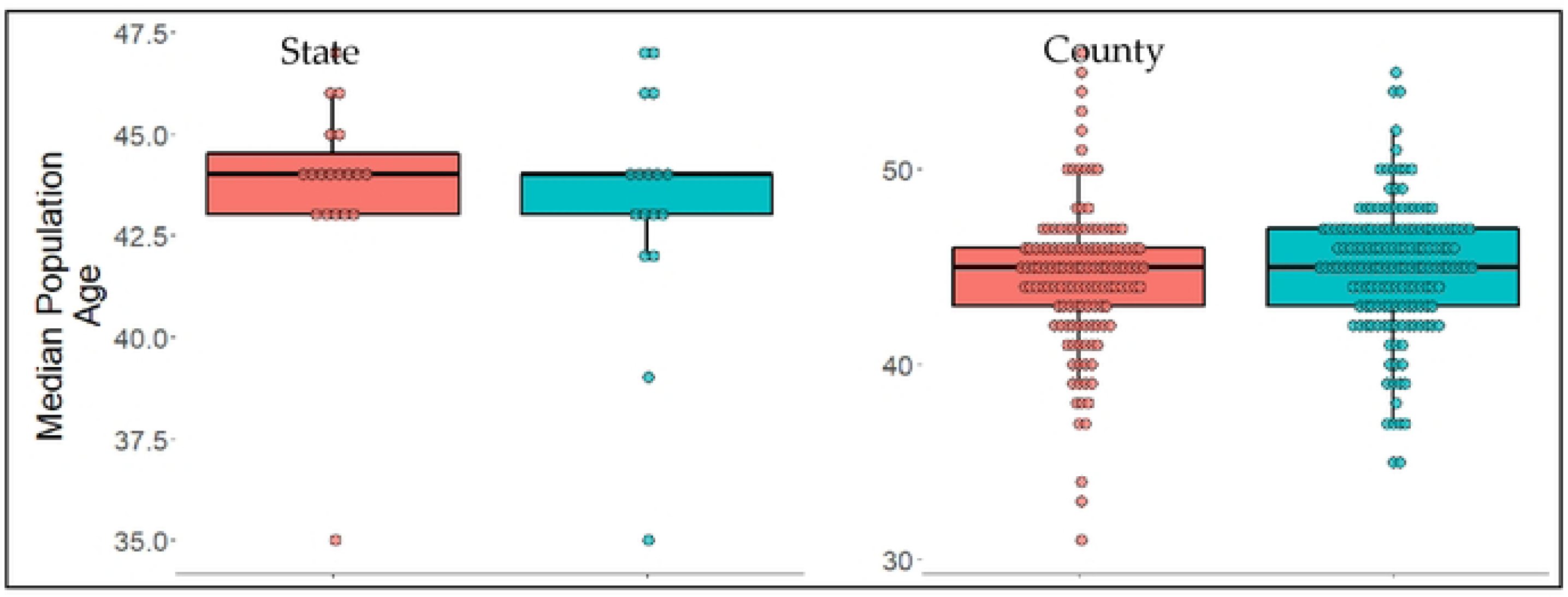
Median age of ACS survey respondents. Non-successful SAHOs are shown on the left in red, successful SAHOs are shown on the right in green.

### • Proportion of eligible population vaccinated

The COVID-19 pandemic has been plagued with mistrust of government officials and the medical community. This mistrust has led to a lack of compliance with public health orders and recommendations, such as SAHOs or receiving vaccinations. Because no quantification of mistrust in government and medical communities exists at the locational specificity needed for this analysis, we utilized the percentage of individuals who had received at least one dose of a COVID vaccine as a proxy for this mistrust. We expected that communities with unsuccessful SAHOs would have lower vaccination rates. To assess this, we tested the null hypothesis that the distribution of vaccination rates as of July 15, 2021, was the same in communities with effective and ineffective SAHOs. These tests yielded non-significant results, and the null hypothesis was not rejected (p=0.850 states; p=0.080 counties). This result can be extrapolated to suggest that mistrust in government and public health systems likely did not influence SAHO effectiveness.

**Figure 10:**
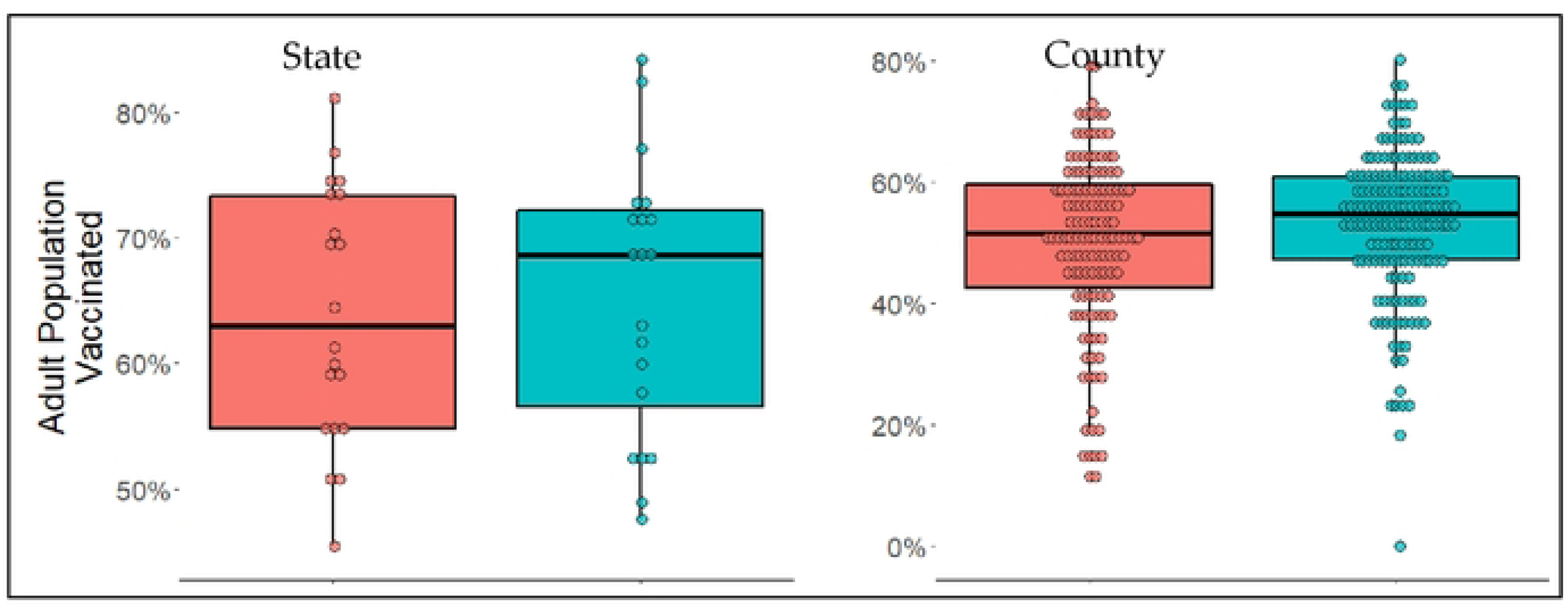
The percent of eligible individuals vaccinated as of July 15, 2021. Unsuccessful SAHOs are shown on the left in red, successful SAHOs are shown on the right in green.

### • Proportion of population who voted for Trump in 2020 presidential elections

Leading up to and during SAHOs in the spring of 2020 there were stark differences in how politicians discussed the threat of COVID-19, and the types of NPI’s they supported. This difference is commonly credited with affecting how populations responded to public health orders. Numerous media articles and scientific studies suggest that individuals who identified as Republican or supported Trump were less likely to adhere to public health orders such as mask mandates and SAHOs. We utilized the voting records from the 2020 presidential election to estimate the proportions of people who supported Trump or the Republican party. As with other variables, we then looked to see if the distribution of the percent of people who voted for Trump was different in communities based on SAHO effectiveness. Surprisingly, a significant difference was not found suggesting that support for Trump did not have a significant influence on SAHO effectiveness (p=0.478 states; p=0.895 counties).

**Figure 11:**
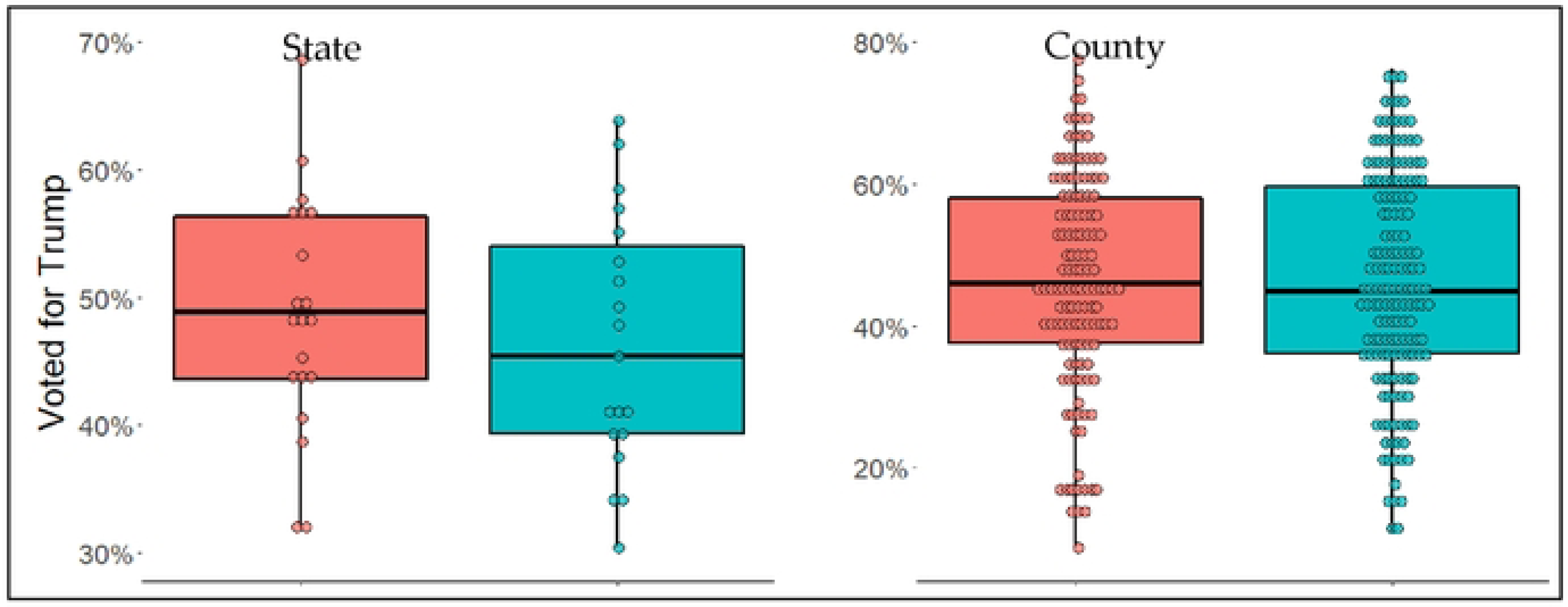
Percent of population that voted for Trump in the 2020 general election. Non-successful SAHOs are shown on the left in red, successful SAHOs are shown on the right in green.

### • Proportion of population who identify as Hispanic and African American

The pandemic has highlighted and deepened the stark inequities that exist along racial and ethnic lines. Studies have shown that COVID-19 has disproportionately affected Black and Hispanic individuals. For example, both Black and Hispanic communities have seen a larger drop in life expectancy during the pandemic than white individuals.^35^ The larger impact of COVID-19 on these communities should not be attributed to racial or ethnic differences but rather the multitude of structural and systemic variables that disadvantage these individuals relative to white individuals. These structural and systemic variables, however, are too numerous and difficult to quantify for this analysis. For this reason, we utilized the percent of a population identifying as African American and the percent of a population identifying as Hispanic as a proxy for the structural and systemic variables which have created a larger burden of COVID-19 in these communities. We tested the null hypothesis that the percentage of individuals identifying as African American or Hispanic in a community was equally distributed between communities with successful and unsuccessful SAHOs. These tests yielded non-significant results, suggesting that whether a SAHO was effective in a community was not associated with the proportion of African American individuals (p=0.573 states; p=0.493 counties) or Hispanic individuals (p=0.950 states; p=0.262 counties) in that community.

**Figure 12:**
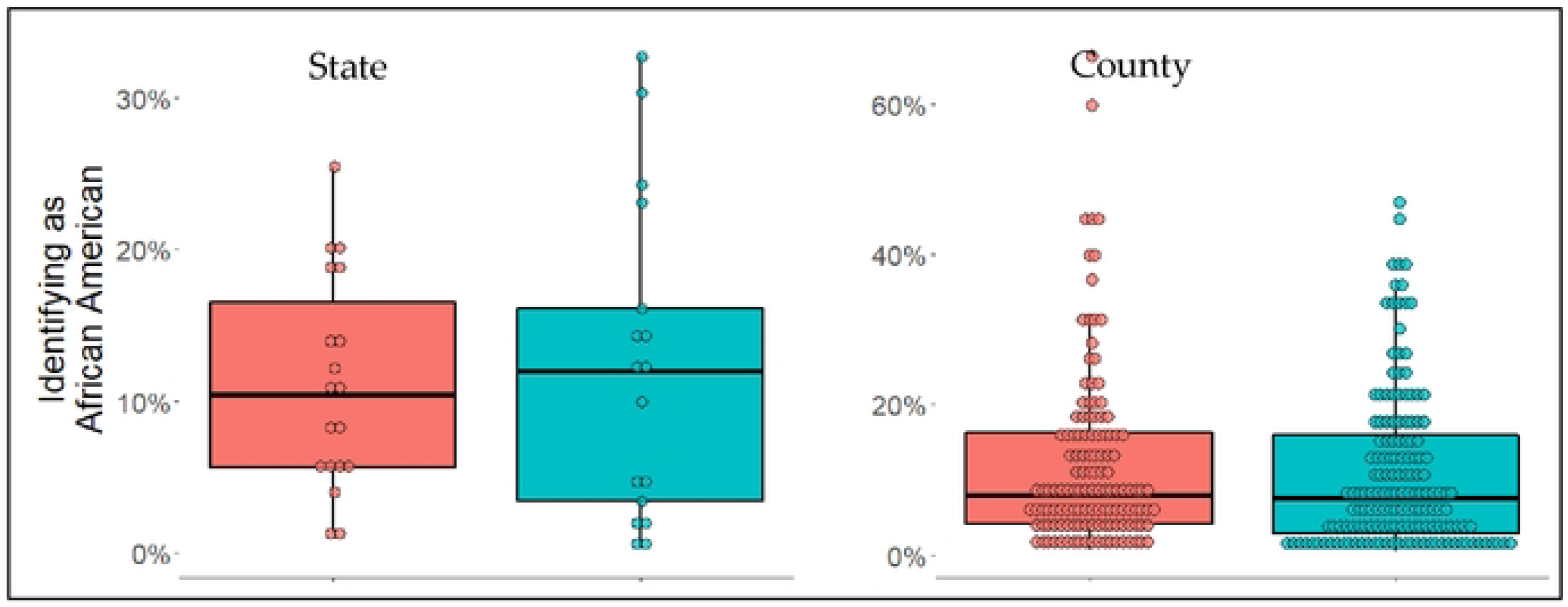
The percent of individuals who identified themselves as African American on the ACS survey. Non-successful SAHOs are shown on the left in red, successful SAHOs are shown on the right in green.

**Figure 13:**
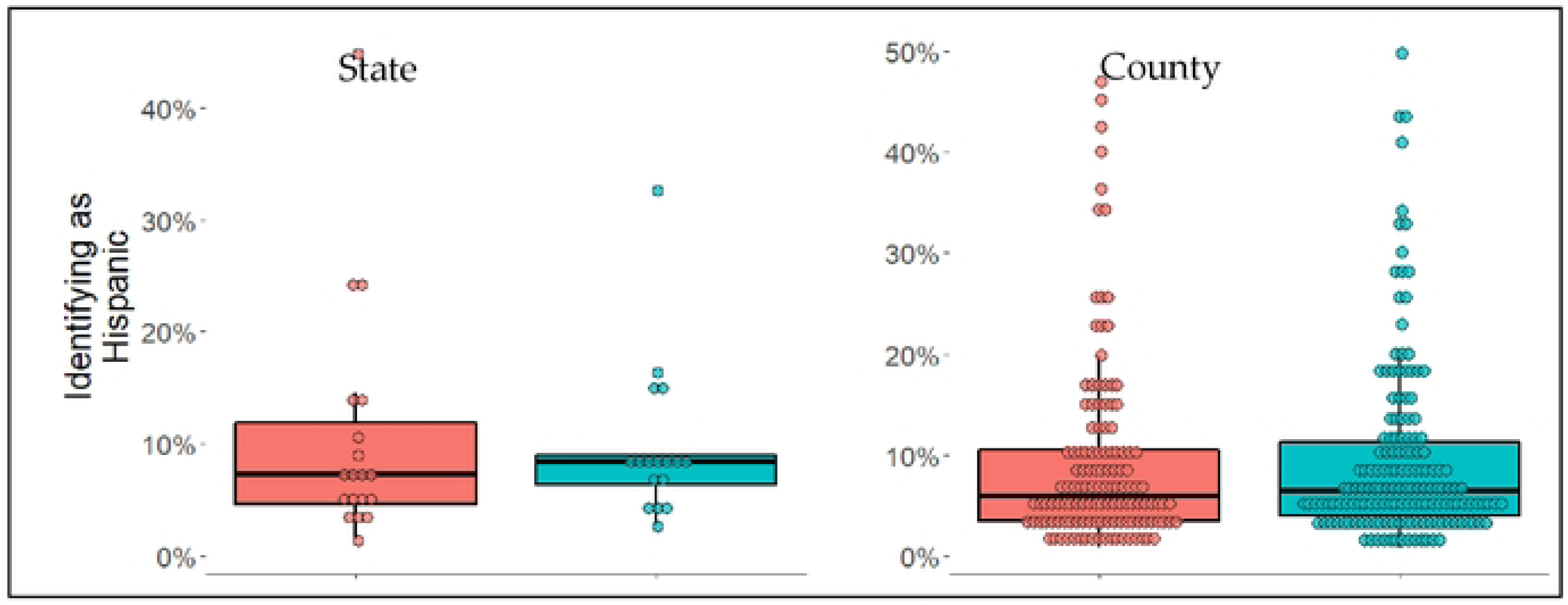
The percent of individuals who identified themselves as Hispanic on the ACS survey. Non-successful SAHOs are shown on the left in red, successful SAHOs are shown on the right in green.

After completing a bivariate analysis of these population characteristics, we conducted a multivariate logistic regression analysis to assess whether some of the variables were confounders or effect modifiers. In R Studio we controlled for each of the 12 variables and found that none of them had more than a 10% impact on any of the other variables. This suggests that there is no confounding or effect modification taking place amongst the variables considered. The cumulative number of days of prior NPI orders remained the only significant result in our multivariate model.

## DISCUSSION

The burden of COVID-19 has varied substantially across the US. Media reports, case studies, and scientific literature, have all commonly pointed to the characteristics of local populations as possible explanations for this variability.^36–38^ Our study aimed to determine which factors may have influenced the effectiveness of SAHOs issued in the U.S. in the spring of 2020. To answer this question, we assessed whether the distribution of 12 characteristics was different in communities with successful versus unsuccessful SAHOs. The characteristics considered were: the time between the first case of COVID-19 and start of a SAHO, the value of R_t_ at start of a SAHO, the cumulative number of days of alternate NPIs prior to the SAHO, the proportion of front-line workers in a community, the educational attainment of a community, the proportion of individuals who lack fluency in English, the median income of a population, the mean age of a population, the proportion of adults who had received at least one dose of a COVID-19 vaccine by July 15^th^ 2021, the percentage that voted for Trump in the 2020 general elections, the proportion of a community that identifies as African American, and the proportion of a community that identifies as Hispanic.

We expected our results to show that SAHOs were more effective in populations with characteristics such as high median income compared to populations with contrasting characteristics such as low median income. This would have been in line with other reports linking population characteristics to variations in the burden of COVID-19. However, we found that the only characteristic with a significant difference was the cumulative number of days of alternate NPI orders prior to a SAHO; this showed that a fewer number of days of prior NPI orders positively correlated with SAHO effectiveness. Interestingly, this result was found at the county but not at the state level. When interpreting this result, it is important to recognize the substantial variation in the data. The county data show that while the medians of the two groups may be different, there are still many effective SAHOs which have a greater number of days of prior NPI orders than ineffective SAHOs. Given the variability in the data it is possible our finding is the result of an opportune sample and not representative of the population. If, however, this finding is representative of the population, a possible explanation for the difference could be fatigue. Counties that had endured a greater number of days of restrictive public health orders prior to SAHO start may have already been stressed or fatigued to the point where complying with SAHOs was not feasible. This reduction in compliance could have caused a reduction in order effectiveness. Another, and perhaps more likely, explanation would be that prior NPI’s minimized the impact of COVID-19 on communities. This could have caused communities with more days of prior NPIs to view SAHOs as unnecessary or an overreaction. It follows that such a perspective could lead populations to be less compliant with a SAHO thereby reducing the order’s effectiveness. This explanation would be in line with other studies that found combinations of less invasive NPIs were almost as effective at reducing spread of COVID as SAHOs.^23,39^

Our findings should encourage those assessing the variable burden of COVID-19 to look beyond the convenient explanations of demographics. Rather they should consider more complex and detailed structural and systemic variables. Our assessment supports the idea that greater in-depth research is necessary to explain the variation in the success of SAHOs. More detailed assessment is particularly important given that polarization of the American public has been increasing and negatively impacting public health.^40^ Inappropriately attributing health outcomes, such as SAHO effectiveness, to factors that contrast communities could deepen the division between them and build further inequity. This would result in undermining the efforts of the public health community.

### Limitations

While our findings highlight the importance of detailed assessment into structural and systemic factors, the study also has significant limitations. This analysis is based on cases reported in the Johns Hopkins University Coronavirus Resource Center Database. While this dataset is thought to be one of the most complete records of COVID-19 cases in the US, it is still far from representing the true number of COVID-19 infections. This is due to a multitude of factors impacting our ability to identify cases throughout the pandemic. Early in the pandemic, a lack of available tests left many people possibly infected with COVID-19 undiagnosed. This effect was compounded by restrictive CDC guidelines which outlined who should get tested for COVID-19. In early 2020 these guidelines restricted testing to individuals with multiple symptoms and travel history to various hot spots such as China, Italy, or Iran.^41^ Tests were also not equally available to all populations, creating testing deserts where fewer cases were detected.^42^ It is also well established that a large proportion of COVID-19 cases are asymptomatic and often undetected. One study found that in New York City, 82%-87% of cases were asymptomatic.^43^ Complicating this further, a higher proportion of asymptomatic cases occur in young and healthy individuals compared to older individuals and people with comorbidities. These factors and others have caused a substantial under-reporting of the number of COVID-19 cases in all datasets, including the one used in this analysis. Studies have attempted to map the proportion of unreported cases across the US.^44^ Such data could be used to adjust case numbers in the Johns Hopkins Database; however, this modification would lack the locational and temporal precision necessary for this analysis. The result of these sources of error in the case data is a measurement bias where cases are ubiquitously under-reported, however, the degree of under-reporting cannot be assumed to be uniform throughout time or between locations.

To further assess the quality of case data, we compared how reported cases in the Johns Hopkins dataset compared to reported cases in the CDC’s data set. Surprisingly we found substantial differences between the two. In some locations there was more than a 20% difference in cumulative cases reported. These differences are thought to stem from methods of data collection. Johns Hopkins actively collects case data from sources, while the CDC passively has public health departments send case data to them. This distinction led us to believe that the Johns Hopkins database is more accurate.

Variations in daily reported cases greatly influence our calculations of the viral reproductive number. The influence of errors in the number of reported cases is especially large when relatively few cases have been reported. Generally, the smaller communities in our sample reported fewer cases which caused them to be disproportionately affected by errors in case numbers. Unfortunately, it is not possible to determine how errors in case reporting influenced R_t_ calculations without knowing how under-reporting varied temporally and geographically.

Another important limitation of our study is that calculations of R_t_ are heavily dependent on the serial interval (SI) of the virus. SI is a measure of how long it takes one infected individual to pass the infection to another person. We used a serial interval of 5.2 days with a standard deviation of 0.7 days which was derived from a meta-analysis.^32^ However, other studies have found that SI varies substantially with location and is also influenced by various public health orders.^45^ For example, a serial interval in a congregate living environment such as a prison would be expected to be lower than the SI for a rural farming community. Similarly, an order prohibiting group gatherings would be expected to increase the SI as the order would reduce the opportunities for individuals to pass viral particles among each other. Knowing this we performed a sensitivity analysis of the SI and its standard deviation. Surprisingly we found that varying the SI by one day from 5.2 to 4.2 or 6.2 altered our values of R_t_ by up to 50%. Varying the SI’s standard deviation by one day also impacted our values of R_t_, in some cases by more than 20%. Notably, these variations also created substantial changes in which communities had effective and ineffective SAHO’s. Published literature on the COVID-19 virus’ reproductive value utilizes a wide range of SI’s which suggests that there may be lack of comparability between these studies, and that reported R values may not be accurate if the SI was not derived from cases geographically and temporally pertinent to the analysis. Numerous studies such as this one, have used a single SI value to derive R values for a range of times and locations. Our findings suggest that these methods are insufficient for generalized analyses, and such reports should be considered with caution. We recommend that future research identifies a more robust method for either calculating R_t_ or qualifying SAHO effectiveness in a manner that is not dependent on SI values. This is especially important given that policy makers have used R values throughout the pandemic to help inform decisions with profound consequences, such as when to end a SAHO.

## CONCLUSION

This analysis is a first look into which factors influence the effectiveness of SAHOs. We found that the cumulative number of days of prior NPI orders may have negatively influenced SAHO effectiveness; however, all other characteristics assessed likely had no impact. It is possible that variability in SAHO effectiveness is derived from systemic (for example access to public health messaging) or structural factors (for example, the financial burden created by a SAHO). It is also possible that the methods used for calculating R_t_ were not sufficiently robust to accurately identify effective and ineffective SAHOs, causing associations to be missed.

If SAHOs become necessary again in the future, policy makers should recognize that the effectiveness of orders issued in spring of 2020 was variable, and that further research is needed to identify the factors responsible for that variability. Given the extreme impact of SAHOs on communities, determining factors that increase or decrease their effectiveness remains imperative. The threat of a future pandemic requiring lockdowns has drawn the attention of the world. Information presented here may help inform how future SAHOs can be implemented in a manner that maximizes their impact while minimizing their duration and harm.

## ACKOWLEDGEMENTS

Dr. Yea-Hung Chen has provided extremely valuable input and insight into how to accomplish the goals of this paper. This project required an enormous amount of data organization and manipulation. Dr. Chen’s guidance was essential to accomplishing this. Additionally, Dr. Chen’s encouragement and enthusiasm for determining factors that impacted performance of communities during the pandemic brought life to the project.

Dr. Brooke Jarvie has contributed to this project substantially by providing feedback, entertaining brainstorming discussions.

Dr. Kelly Sanders has also been a consistent supporter of this project. Her feedback on, and enthusiasm for this project has greatly improved it. Dr. Sanders’ willingness to invest time into this project has been tremendously appreciated and valued.

## Data Availability

All relevant data will be in supporting information files

